# Reduced levels of convalescent neutralizing antibodies against SARS-CoV-2 B.1+L249S+E484K lineage

**DOI:** 10.1101/2021.09.13.21263430

**Authors:** Diego A. Álvarez-Díaz, Katherine Laiton-Donato, Orlando Alfredo Torres-García, Hector Alejandro Ruiz-Moreno, Carlos Franco-Muñoz, Maria Angie Beltran, Marcela Mercado-Reyes, Miguel Germán Rueda, Ana Luisa Muñoz

**Author notes:** These authors contributed equally. **Corresponding author** Ana L Muñoz, Calle 23 #116-31, Bodega 26, Bogotá, Colombia.

## Abstract

The E484K mutation at the SARS-CoV-2 Spike protein emerged independently in different variants around the world, probably as part of the ongoing adaptation of the virus to the human host, and has been widely associated with immune escape from neutralizing antibodies generated during previous infection or vaccination. In this work, the B.1+L249S+E484K lineage was isolated along with A.1, B.1.420 and B.1.111 SARS-CoV-2 lineages without the E484K mutation and the neutralizing titer of convalescent sera was compared using microneutralization assays. While no significant differences in the neutralizing antibody titers were found between A1 and B lineages without the E484K mutation, the neutralizing titers against B.1+L249S+E484K were 1.5, 1.9, 2.1, and 1.3-fold lower than against A.1, B.1.420, B.1.111-I, and B.1.111-II, respectively. However, molecular epidemiological data indicate that there is no increase in the transmissibility rate associated with this new lineage. Hence, although the evidence provided in this study support a Variant of Interest Status (VOI) for the B1+L249S+E484K lineage, enhanced laboratory characterization of this particular lineage and other emerging lineages with the E484K mutation should be carried out in individuals with immunity acquired by natural infection and vaccination. This study accentuated the capability of new variants with the E484K mutation to be resistant to neutralization by humoral immunity, and therefore the need to intensify surveillance programs.

**HIGHLIGHT:**

⍰ The E484K mutation in B.1+L249S+E484K appears not to affect the viral titer
⍰ Sensitivity of lineages without E484K mutation to neutralizing antibodies did not change
⍰ B.1+L249S+E484K lineage shows a reduction in its neutralizing capacity

## 1. INTRODUCTION

The pandemic of coronavirus disease 2019 (COVID-19) caused by severe acute respiratory syndrome coronavirus 2 (SARS-CoV-2) is rapidly changing. It is known that, following the infection, a polyclonal set of antibodies against SARS-CoV-2 proteins are produced, playing a crucial role in the acquired immunity against this virus. In particular, antibodies generated against the spike (S) protein are well known for their ability to compromise the host cell receptor binding and membrane fusion. This leads to blocking the entry of the virus into the host cell, and these antibodies are called neutralizing antibodies (1).

However, the adaptive process of SARS-CoV-2 to human host has resulted in the emergence of antigenically distinct and/or more virulent variants with evidence of reduced neutralization by antibodies generated from natural infection or vaccination; a matter of great importance in the process of selecting the antigenic component in vaccine formulation (2).

Whereby, the US government interagency group and Centers for Disease Control and Prevention (CDC), proposed a hierarchical variant classification scheme with three classes of SARS-CoV-2 variants, namely, Variant of Interest (VOI), Variant of Concern (VOC) and, Variant of High Consequence (VOHC). Here, the presence of substitutions or combinations of substitutions at the Spike (S) protein, associated with reduced neutralization by antibodies generated from previous infection or vaccination, is one of the critical attributes to classify as VOI. The escalation to VOC depends on scientific evidence such as laboratory confirmation of significant escape to neutralizing antibodies and/or epidemiological data supporting increased transmissibility and disease severity. Finally, although no VOHC have been identified so far, it is expected that a VOHC has clear evidence of diagnostic failure, a significant reduction in vaccine effectiveness, therapeutics, and more severe clinical disease (3).

In February 2020, in the early stages of the pandemic, the B.1 lineage emerged in Europe with the characteristic D614G mutation in the S protein, which distinguishes it from the A and B ancestral lineages (4). This lineage rapidly became the most prevalent (5), even in South America, where the first introduced cases corresponded to the A.1 lineage with the conserved D614 position (6), and several studies provided evidence on the association of this mutation with fitness advantages without significant impacts on the severity of infection or neutralizing antibody titers (4,7,8).

Later, the E484K mutation in the S protein emerged independently in different VOI and VOC, probably by evolutionary convergence (9,10) and was associated with reduced antibody neutralization (11), antiviral drug resistance (12) and a slightly enhancing of ACE2 affinity (13). In fact, the E484K mutation is a significant genetic marker, and its presence is considered enough to qualify a variant for VOI status (14).

In line with this, SARS-CoV-2 genomic surveillance at Colombia’s National Institute of Health (INS), identified a highly divergent SARS-CoV-2 lineage characterized by the presence of 21 substitutions, including two amino acid changes in the S protein (L249S and E484K). For this reason, it was proposed for lineage reassignment (B.1+L249S+E484K) and laboratory evaluation of neutralizing antibodies (15).

Several methods have been documented for the assessment of neutralizing antibodies against SARS-CoV-2, among them, pseudovirus-based protocols have proved well correlation with neutralization titers (16–19), however, the gold standard is the *in vitro* neutralization using replication-competent virus because their sensitivity is higher and allow the evaluation of aspects such as viral fitness and the impact of mutations in proteins other than Spike (20,21).

Thus, in this work, we determined the neutralizing antibody titers in convalescent sera against B.1+L249S+E484K and three lineages (A.1, B.1.420, and B.1.111) without the E484K mutation using microneutralization assays to evaluate the potential impact of the E484K mutation in this new lineage on the sensitivity to convalescent neutralizing antibodies.

## 2. MATERIAL AND METHODS

### 2.1 Human subject collection

The samples were collected between March 2020 and February 2021; all subjects enrolled in this research responded voluntarily to an informed consent formulary previously approved by the Ethics Committee of Colombian National Health Institute (CEMIN)-10-2020. This study was conducted in compliance with ethical principles of the Declaration of Helsinki and to the conditions provided by the Ministry of Health - Colombia.

### 2.2 Cells

African green monkey kidney Vero E6 cells (ATCC CRL-1586™) were used to propagate the SARS-CoV-2 isolates and the neutralization assays. Cells were cultured in Dulbecco’s Modified Eagle’s medium (DMEM) (Lonza®, Catalog No. 12-604Q) supplemented with 10% (v/v) heat-inactivated fetal bovine serum (FBS) (Biowest®, Catalog No. S18b-500), and 100 U/mL penicillin and streptomycin (Lonza®, Catalog No. 17-602F) at 37°C with 5% CO_2_.

### 2.3 Sample selection and virus isolation

Nasopharyngeal swab specimens from volunteer participants from the five Colombian regions were collected based on the representativeness and virologic criteria, following the Pan American Health Organization (PAHO) guidance for SARS-CoV-2 samples selection (22). Samples with positive real-time RT-PCR for SARS-CoV-2 and PANGO lineage assignment (23), following the nanopore ARTIC network protocol (24), were selected for virus isolation in Vero E6 monolayers.

For this, each sample was diluted 1:2 with DMEM supplemented with 2% FBS. The dilutions were filtered through a 0.2um membrane and used to inoculate 7.5 x10^5^ Vero E6 cells seeded the previous day in T-25 flasks. The inoculum was incubated for 1 hour at 37 °C in a 5% CO_2_ environment. Finally, 4 mL of DMEM medium supplemented with 4% FBS were added, and virus-induced cytopathic effect (CPE) was examined daily for up to 7 days. When CPE was observed, culture supernatant was collected and centrifuged at 300xg for 5 min at room temperature, distributed in aliquots of 500uL, and stored in liquid nitrogen (25). All the procedures handling the infected cell cultures were held in a biocontainment laboratory.

### 2.4 Phylogenetic analysis

We recovered 1856 sequences from SARS-CoV-2 infections in Colombia from the GISAID database. The sequence dataset was aligned using the MAFFT software v7 (26,27). The alignment was manually curated to remove UTRs and correct possible misalignments. Sequences with genome coverage lower than 90 percent were removed, as well as redundant identical sequences. The final aligned dataset contained 400 sequences with representatives from each lineage circulating in Colombia by July 2021 in each region. A maximum likelihood tree reconstruction was performed with the GTR+F+I+G4 nucleotide substitution model using IQTREE. The substitution model was selected according to the lowest BIC score using IQTREE modelfinder (26). Branch support was estimated with 1000 replicates of an SH-like approximate likelihood ratio test (SH-aLRT), and 1000 ultrafast bootstrap replicates.

### 2.5 Virus titration

Virus titers were determined by the Reed and Muench tissue culture infective dose (TCID_50_) endpoint method (28), after three independent assays. Briefly, for titration, 1.5 × 10^5^ Vero E6 cells/mL were plated into 96-well plates 16 to 24 hours before infection. Then, the virus stock was diluted serially from 10−1 to 10−10, and 100 µL of each dilution was added to the respective well (Eight replicates were performed for each dilution) and incubated in 5% CO_2_ at 37 I]C for 72 h. Finally, the TCID_50_ endpoint was determined.

### 2.6 Microneutralization and binding antibody assays

Serum samples were obtained from five convalescent donors with COVID-19 diagnosis confirmed by real time RT-PCR on nasopharyngeal swab specimens at least 15 days before inclusion. A serum bank specimen from 2019 was used as a pre-pandemic negative control. Additionally, SARS-CoV2 IgG anti-Nucleocapsid antibodies detection on the Allinity ci series (Abbott, Chicago,IL, USA) was done as screening, where relative light units (RLU) >1.4 were interpreted as positive. Serum samples were conserved at -20°C until the moment of the analysis.

The microneutralization assay was adapted from the methodology published by Algaissi A and Hashem A (25). Briefly, the presence of neutralizing antibodies was assessed by means of eight 2-fold dilutions (1:20 to 1:2560) of heated inactivated serum samples tested against 2000 TCID_50_ of each variant virus stock in supplemented DMEM with 2% FBS for one h at 37 ⍰C. After that, the suspension was transferred on a monolayer of 1.5 × 10^4^ Vero E6 and incubated at 37°C with 5% CO_2_ for 3 days. The neutralizing titer was determined as the highest serum dilution at which no CPE was observed under inverted microscope Primovert (Zeiss). Each sample was tested five times, in three independent tests.

### 2.7 Statistical analysis

For analysis of differences in serum neutralizing antibody titers, data were log transformed and one-way ANOVA using the DMS multiple comparison correction was used for columnated data while two-way ANOVA using the Tukey multiple comparison correction was used for grouped data. Results with p values <0.05 were considered significant. The data were analyzed in GraphPad Prism (v9.0.2).

## 3. RESULTS

### 3.1 Successful isolation of SARS-CoV-2 lineages with and without the Spike E484K mutation

Five SARS-CoV-2 isolates representing four different PANGO lineages were selected for the MN assays (Table 1). Remarkably, the isolate EPI_ISL_1092007 currently assigned to the B.1 lineage (PANGO v.3.1.7 2021-07-09) was proposed for lineage reassignment (B.1+L249S+E484K) and laboratory evaluation of neutralizing antibodies in convalescent sera (15). The remaining isolates represented SARS-CoV-2 lineages without the E484K mutation as follows; EPI_ISL_49816 representing the A1 lineage, it was the only isolate without the characteristic D614G mutation in the S protein obtained in this study. The other isolates correspond to the B.1.420 (EPI_ISL_52696 B.1.420) and B.1.111 (EPI_ISL_526971 and EPI_ISL_794659) lineages. Regarding B.1.111 isolates, although both share the mutation pattern characteristic of the B.1.111 lineage (Spike D614G, NS3 Q57H and NSP12 P323L), several divergent genome wide mutations were observed between those isolates, for example, EPI_ISL_794659 have two additional mutations at the S protein (Spike T859I and W152R) (Table 1).

**Table 1.** Demographic data and genomic characteristics of the viral isolates selected for MN assays.

| Pango Lineage* | Isolate name (GISAID AC. No.) | Colombian Region | AA Substitutions | Disease progression - Outcome | Patient status | Symptoms |
| --- | --- | --- | --- | --- | --- | --- |
| A.1 | EPI_ISL_498169 | Andean | NS8 L84S, NSP1 G105S | Symptomatic | Live | Odynophagia |
| B.1.420 | EPI_ISL_526969 | Bogotá DC | Spike D614G, N A119T, NSP12 A43S, NSP12 P323L | Symptomatic | Live | Cough |
| B.1.111-I | EPI_ISL_526971 | Bogotá DC | Spike D614G, N M234I, NS3 Q57H, NSP3 S1285F, NSP12 P323L | Asymptomatic | Live | None |
| B.1.111-II | EPI_ISL_794659 | Andean | Spike D614G, Spike T859I, Spike W152R, N T205I, NS3 Q57H, NS3 T223I, NSP1 I114T, NSP5 K90R, NSP6 A136V, NSP12 P323L | Hospitalized | Live | Fever, headache |
| B.1 (B1.111+E484K) | EPI_ISL_1092007 | Caribbean | Spike D614G, Spike E484K, Spike L249S, N T205I, NS3 Q57H, NS6 I18V, NS7a L17F, NS7b A15S, NSP5 V157L, NSP12 P323L, NSP13 L325F, NSP13 T153I, NSP15 V314F, NSP16 G213N | No Data | Live | No data available |

Viral stocks of the five isolates titrated by the Reed and Muench method yielded similar titers reaching 10^5^ TCID_50_/mL (Table 1). Thus, the E484K mutation in B.1+L249S+E484K appears not to affect the viral titer in Vero E6 cells (Table 2) or the virus ability to induce a cytopathic effect (Fig. 1A).

**Table 2.** Viral titers of SARS-CoV-2 isolates estimated by the Reed and Muench method.

| Lineage | Titer (TCID<sub>50</sub> /mL) |
| --- | --- |
| A.1 | 3.91 x10<sup>5</sup> TCID<sub>50</sub> /mL |
| B.1.420 | 3.0 x10<sup>5</sup> TCID<sub>50</sub> /mL |
| B.1.111-I | 3.2 x10<sup>5</sup> TCID<sub>50</sub> /mL |
| B.1.111-II | 4.67 x10<sup>5</sup> TCID<sub>50</sub> /mL |
| B.1+L249S+E484K | 4.53 x10<sup>5</sup> TCID<sub>50</sub> /mL |

**Figure 1.**
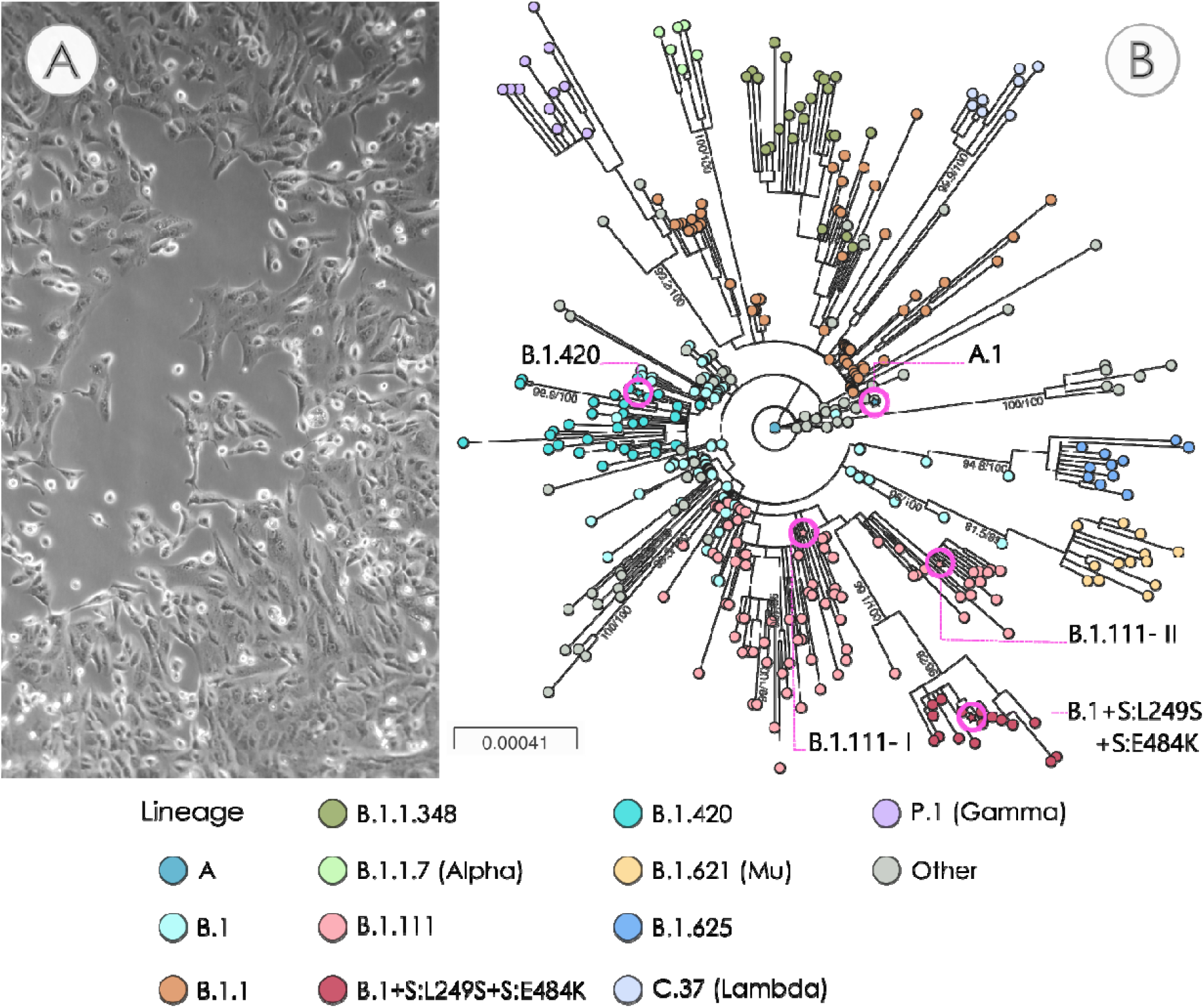
Phylogeny of SARS-CoV-2 lineages selected for the MN assays. A.) Representative image of Vero E6 cells infected with SARS-CoV-2. B.) Maximum likelihood phylogeny of SARS-CoV-2 representative lineages by July 2021. The tree was reconstructed by maximum likelihood with the estimated GTR+F+I+G4 nucleotide substitution model for the dataset of 400 genomes. The scale represents nucleotide substitutions per site. The interactive phylogeny and map are available at https://microreact.org/project/fNyvqQHycKrwPjKFMDo6xD/48df9688

### 3.2 Phylogenetic analysis reveals B.1.111 sublineages

Given the close lineage assignment of four isolates as lineage B.1 or sublineages B.1.111 and B.1.420, we performed a maximum likelihood phylogenetic analysis to confirm their lineage and placement within a tree with 400 representative sequences from each lineage circulating in Colombia by July 2021 (Fig. 1B). The lineage assignment of the five isolates was confirmed in the phylogenetic reconstruction. The isolates B.1.111-I and B.1.111-II share a lineage designation but different mutation profiles (Table 1). The major number of mutations in B.1.111-II indicates a greater divergence consistent with the sampling date and the expected accumulation of mutations during the pandemic, as observed in the tree (Fig. 1B). The isolate B.1.111-I is located close to the node defining the B.1.111 lineage, while B.1.111-II is placed in a monophyletic group along with more B.1.111 sequences containing the Spike mutation W152R, which is not as divergent as B.1+L249S+E484K. Furthermore, in line with the previous report, the B.1+L249S+E484K sample genome is grouped in a sublineage of the B.1.111 lineage (15).

### 3.3 Reduced neutralization antibody titers against B1+L249S+E484K in convalescent sera

We initially characterized five convalescent COVID-19 patients previously diagnosed with SARS-CoV-2 infection by RT-qPCR, sampled at a median of 90d (range 30–150d) post- onset of symptoms, using SARS-CoV2 IgG anti-Nucleocapsid antibodies detection kit (Abbott, Chicago,IL, USA). All individuals demonstrated greater reactivity to nucleocapsid protein than pre-pandemic control, which was comparable to the obtained MN titers (Table 3). Thus, neutralizing antibodies correlated with nucleocapsid binding antibodies (Fig. 2C).

**Table 3.** Clinical data of convalescent subjects.

| Patient Number | Symptoms | IgG (RLU) | MN titer against Lineage B.1.111-I |
| --- | --- | --- | --- |
| 1 | NA | 0.275 | 0 |
| 2 | Adynamia, cough, rhinorrhea breathing difficulties | 3.64 | 1:80 |
| 3 | anosmia and odynophagia | 5.91 | 1:80 |
| 4 | headache, adynamia, fever and rhinorrhea, ageusia | 3.26 | 1:80 |
| 5 | adynamia, rhinorrhea | 6.31 | 1:320 |
| 6 | cough, fever and odynophagia | 3.596 | 1:160 |

**Figure 2.**
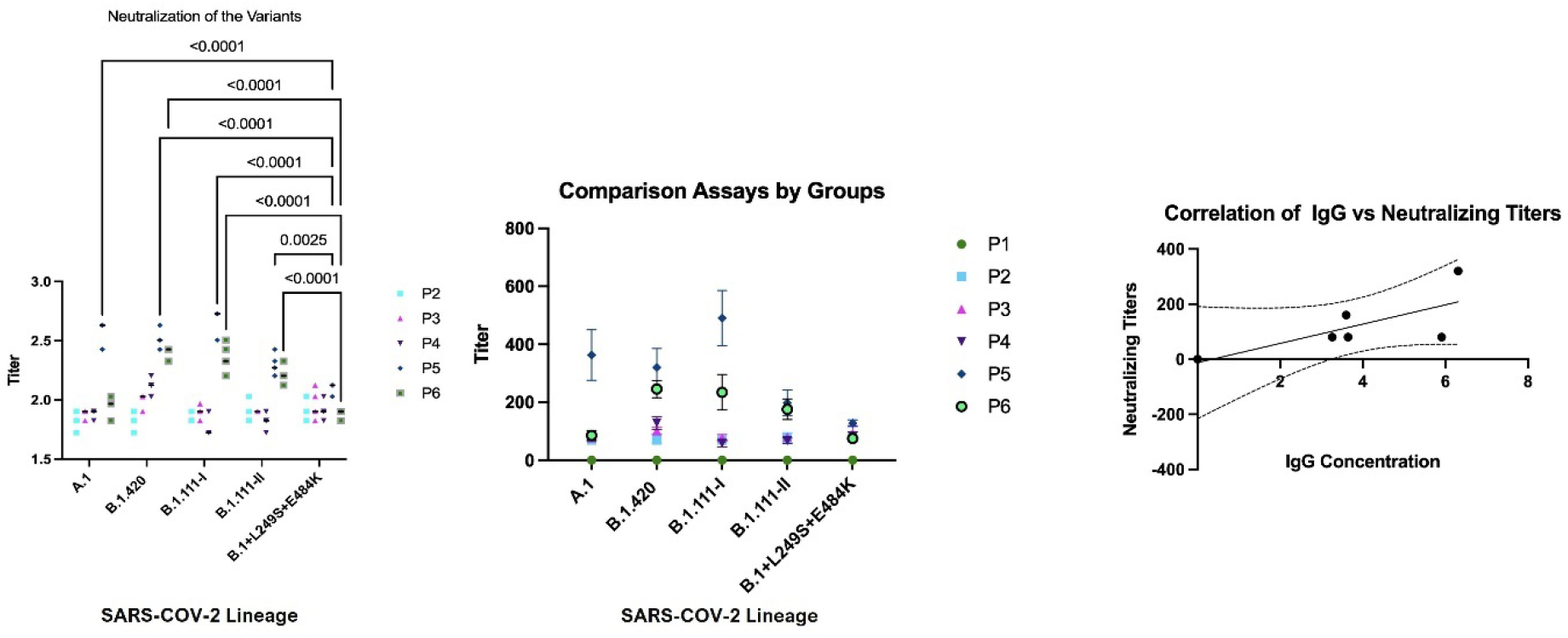
Neutralizing titers of convalescent sera against SARS-CoV-2 A.1, B.1.420, B.1.111, and B.1+L249S+E484K lineages. Panel A. Correlation of neutralizing titers against B.1+L249S+E484K in relation to SARS-CoV-2 lineages. Panel B. Neutralizing titers of convalescent sera against SARS-CoV-2 A.1, B.1.420, B.1.111, and B.1+L249S+E484K lineages. Panel C. B1.111-I sublineage neutralization among convalescent COVID-19 patients and its correlation with SARS-CoV2 IgG anti-Nucleocapsid antibody titers

Then, the neutralizing capacity of these sera was evaluated using eight 2-fold serially diluted sera (1:20 to 1:12560) against A.1, B.1.420, B.1.111, and B.1+L249S+E484K lineages to determine the MN50 titer of each serum sample. Results from three independent assays evidenced differences between the neutralization titers obtained against the five isolates. Specifically, the neutralizing titers against B.1+L249S+E484K were 1.5, 1.9, 2.1 and 1.3-fold lower than against A.1, B.1.420, B.1.111-I and B.1.111-II, respectively (Fig. 2A). Furthermore, the neutralizing titers for the B.1.111-II variant were 1.6-fold lower in relation to the variant B.1.111.I (P< 0.0001) (Fig. 2B). Finally, no significant differences between neutralizing antibody titers were observed between A1 and B lineages without E484K mutation (Figure 2B).

### 3.4 Molecular epidemiological data support a decrease of B.1+L249S+E484K cases between March-2020 and July-2021

The spatiotemporal distribution pattern of the most representative SARS-CoV-2 lineages in Colombia between March 2020 and July 2021 shows a significant country level decrease of cases associated with the lineages evaluated in this study (Fig. 3). By January 2021, B.1.111 was predominant and widely distributed across the five Colombian regions, followed by B.1 and B.1.420 (Fig. 3A and 3B). This scenario changed by July 2021, with a notable decrease in these lineages by two-to three-fold, which were displaced by P.1 and B.1.621. Remarkably, B.1+L249S+E484K was exclusively distributed in the Caribbean region between March-2020 and July-2021, and the decline of cases by the end of this period suggests a limited community transmission (Fig. 3A and 3B).

**Figure 3.**
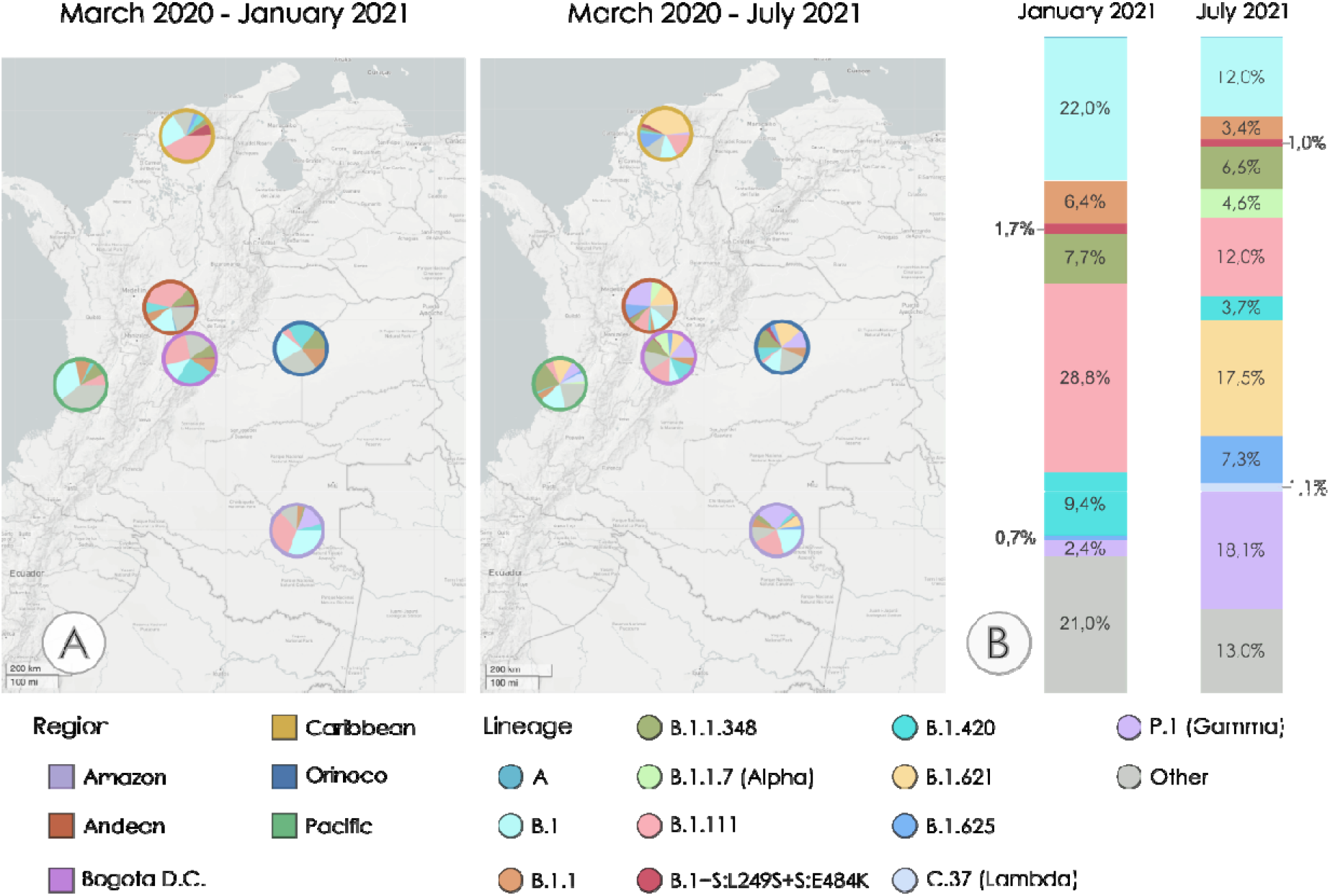
Spatiotemporal distribution of the most representative SARS-CoV-2 lineages in Colombia between January 20210 and July 2021. A.) SARS-CoV-2 lineage distribution in the five Colombian regions and Bogotá D.C March 2020 - January 20210 (left), and March-July 2021 (right). Ring colors represent the region. Interactive map available at https://microreact.org/project/sZ7jeqhSJ3bihFGBE1TvVG/0e4c2d4d. Country level SARS-CoV-2 percentages up to January 2021 (left), and up to July 2021 (right).

## 4. DISCUSSION

The ongoing global surveillance programs have revealed the emergence of variants harboring mutations in Spike, the principal target of neutralizing antibodies. This study shows the neutralizing activity of natural infection-elicited antibodies against four SARS-CoV-2 lineages, including B.1+L249S+E484K.

The E484K substitution, located at the receptor-binding domain (RBD), is continuously and independently occurring in emerging SARS-CoV-2 VOCs and VOIs across all over the world (3). This phenomenon is probably an example of selective immune pressure and receptor-binding affinity optimization as part of the ongoing adaptation of the virus to the human host (9,29).

Our results suggest that E484K mutation in B.1+L249S+E484K does not affect the viral titer, however it reduces antibody neutralization, when compared to the A.1, B.1.420, B.1.111 lineages, all of them without the E484K mutation. In agreement with previous reports, the single E484K mutation was associated with reduced neutralizing activity of convalescent and post-vaccination (Pfizer–BioNTech) sera, against replication-competent SARS-CoV-2, and pseudoviruses (11). Moreover, the resistance to neutralizing antibodies in other lineages carrying this mutation has been described in B.1.1.7, B.1.351, and P.1 (9,30). It is essential to clarify that it has been reported that variants with the E484K mutation have no alterations in its stability or in host cell entry (31).

We also observed a slightly higher neutralizing antibody titer against B.1.111-I compared with B.1.111-II (Fig. 2); this could be explained by the presence of two additional mutations in the S protein, T859I and W152R (Table 1), because the presence of mutations at the same positions have been reported independently in the VOIs B.1.526 (Iota) (32) and B.1.429 (33), respectively.

Remarkably, the phylogenetic maximum likelihood phylogenetic tree reconstructed in this study grouped the isolates B.1+L249S+E484K and B.1.111-II in the same clade (Fig. 1B), which evidences the importance of phylogenetic analysis in understanding phylogenetic relationships beyond the PANGOLIN assignment in the face of the emergence of variants with particular mutations.

While the MN assays unequivocally evidenced reduced neutralization antibody titers against B1+L249S+E484K in convalescent sera (Fig. 2A), molecular epidemiological data indicate that there is no increase in the transmissibility rate associated with this new lineage (Figure 3). Hence, the B1+L249S+E484K lineage must be regarded as a VOI at least for Colombia, furthermore, surveillance of this particular lineage and other emerging lineages with the E484K mutation should be carried out in individuals with immunity acquired by natural infection and vaccination.

A limitation of this study is the small number of convalescent samples tested, and the absence of samples from vaccinated individuals, future work with more subjects, including vaccinated individuals will help to determine the clinical impact of this mutation, as well as its role in the effectiveness of currently approved vaccine strategies.

## 5. CONCLUSION

These results suggest the emergence of a new SARS-CoV-2 lineage with the ability to escape from neutralizing humoral immunity. As the virus continues to adapt to the human host, the accumulation of these mutations on aspects such as the immune response against natural infection or vaccination is unknown. Consequently, it is necessary the intensification of the genomic surveillance programs and the refinement of protocols for the evaluation of point and multiple mutations and their association with the escape from neutralizing antibodies.

## Data Availability

The authors confirm that the data supporting the findings of this study are available within the article [and/or] its supplementary material.

## ACKNOWLEDGEMENTS

Authors gratefully acknowledge Rotary International for equipment’s donation. Authors also acknowledge all the contributing scientists publishing viral sequences in GISAID database, as well as its administrators for supporting rapid sharing of genomic data.

## AUTHORS’ CONTRIBUTIONS

**Diego A. Álvarez-Díaz:** Conceptualization, Methodology, Investigation, Formal analysis, Data Curation, Writing - Original Draft. **Katherine Laiton-Donato**: Investigation, Data Curation, Formal analysis. **Orlando Alfredo Torres-García**: Validation, Formal analysis, Data Curation. **Carlos Franco-Muñoz**: Data Curation. **Hector Alejandro Ruiz**: Software, Formal analysis, Writing - Original Draft. Maria Angie Beltran: Investigation. **Marcela Mercado-Reyes**: Conceptualization, Methodology, Funding acquisition, Supervision. **Miguel Germán Rueda**: Conceptualization, Methodology, Funding acquisition, Supervision. **Ana Luisa Muñoz**: Conceptualization, Methodology, Investigation, Formal analysis, Data Curation, Writing - Original Draft.

